# Determination of daily reproduction numbers of SARS-CoV2 based on death cases suggests more rapid initial spread in Italy and the United States

**DOI:** 10.1101/2020.03.28.20046094

**Authors:** Armin Ensser, Klaus Überla

## Abstract

Population density, behaviour and cultural habits strongly influence the spread of pathogens. Consequently, key epidemiological parameters may vary from country to country. Confirmed COVID-19 cases in in China have been used to estimate those parameters, that vary largely (reviewed in 1). The estimates also depend on testing frequency and case definitions that are prone to change during ongoing epidemics, providing additional uncertainties. The rise in fatal cases due to SARS-CoV2 could be a more reliable parameter, since missing of deaths is less likely. In the absence of changes in the management of severe COVID-19 cases, the rise in death cases should be proportional to the rise in virus infections. Although the fluctuating low numbers of fatal cases very early in the epidemic may lead to some uncertainty, more than 100 deaths per day are reported since 10.03.2020 in Italy and since 21.03.2020 in the US. Therefore, the dynamics of deaths were analysed to estimate the daily reproduction numbers (Rt) and the effectiveness of control measures.

Thus, our analysis provides evidence that basic epidemiological parameters differ between countries to an extent compromising epidemiological predictions of the pandemic. It also suggests that suppression of spread in Italy and the US may be more difficult to achieve. Although we assume that variations in social behaviour are responsible for the different estimates of R0, selection of more rapidly spreading variants of SARS-CoV-2 cannot be excluded. Despite uncertainty in the reliability of the data used and lack of information on possible changes in the effectiveness of registration of COVID-19 deaths during the observation period, our findings should be considered as a working hypothesis demanding further investigations. As the number of deaths rapidly increases worldwide, we encourage more sophisticated modelling of the epidemic based on the dynamics of death cases by experts in the field.

---

Population density, behaviour and cultural habits strongly influence the spread of pathogens. Consequently, key epidemiological parameters may vary from country to country. Confirmed COVID-19 cases in in China have been used to estimate those parameters, that vary largely (reviewed in ^1^). The estimates also depend on testing frequency and case definitions that are prone to change during ongoing epidemics, providing additional uncertainties. The rise in fatal cases due to SARS-CoV2 could be a more reliable parameter, since missing of deaths is less likely. In the absence of changes in the management of severe COVID-19 cases, the rise in death cases should be proportional to the rise in virus infections. Although the fluctuating low numbers of fatal cases very early in the epidemic may lead to some uncertainty, more than 100 deaths per day are reported since 10.03.2020 in Italy and since 21.03.2020 in the US. Therefore, the dynamics of deaths were analysed to estimate the daily reproduction numbers (R_t_) and the effectiveness of control measures.

Daily death cases from 21.2.2020 to 27.03.2020 were downloaded from ECDC ^2^. A three day sliding period was used to smoothen day to day variations. Fold increases after 7 days were determined for each day. Assuming a serial interval of 4 days ^3,4^ daily reproduction numbers (R_t_) were calculated (Fig. 1A). For Italy, this resulted in mean R_t_ values of approximately 3.4 between February 22^nd^ and March 1^st^. Virus is estimated to be acquired approximately 19 to 29 days before the day of death assuming 4 to 7 days of mean incubation period ^5^ and 15 to 22 days from onset of symptoms to death^6^. Thus the R_t_ values plotted from March 2^nd^ to March 12^th^ are likely due to infections occurring between February 2^nd^ and February 22^nd^. The first confirmed Italian cluster of COVID19 dates to February 22^nd^ indicating that the R_t_ of 3.4 determined for deaths from February, 22nd to March 1st represents the basic reproduction number, R_0_, for SARS-CoV2 in Italy. Increasing awareness of SARS-CoV-2 spread and obligatory social distancing measures progressively introduced starting February 22^nd^ are most likely responsible for the continuous decline of the R_t_ values derived from deaths occurring between March 2nd and March 12th. An average age of fatal cases of 81 years ^7^ may have led to more rapid progression to death explaining a faster decline of R_t_ values than expected.

**Figure 1.**
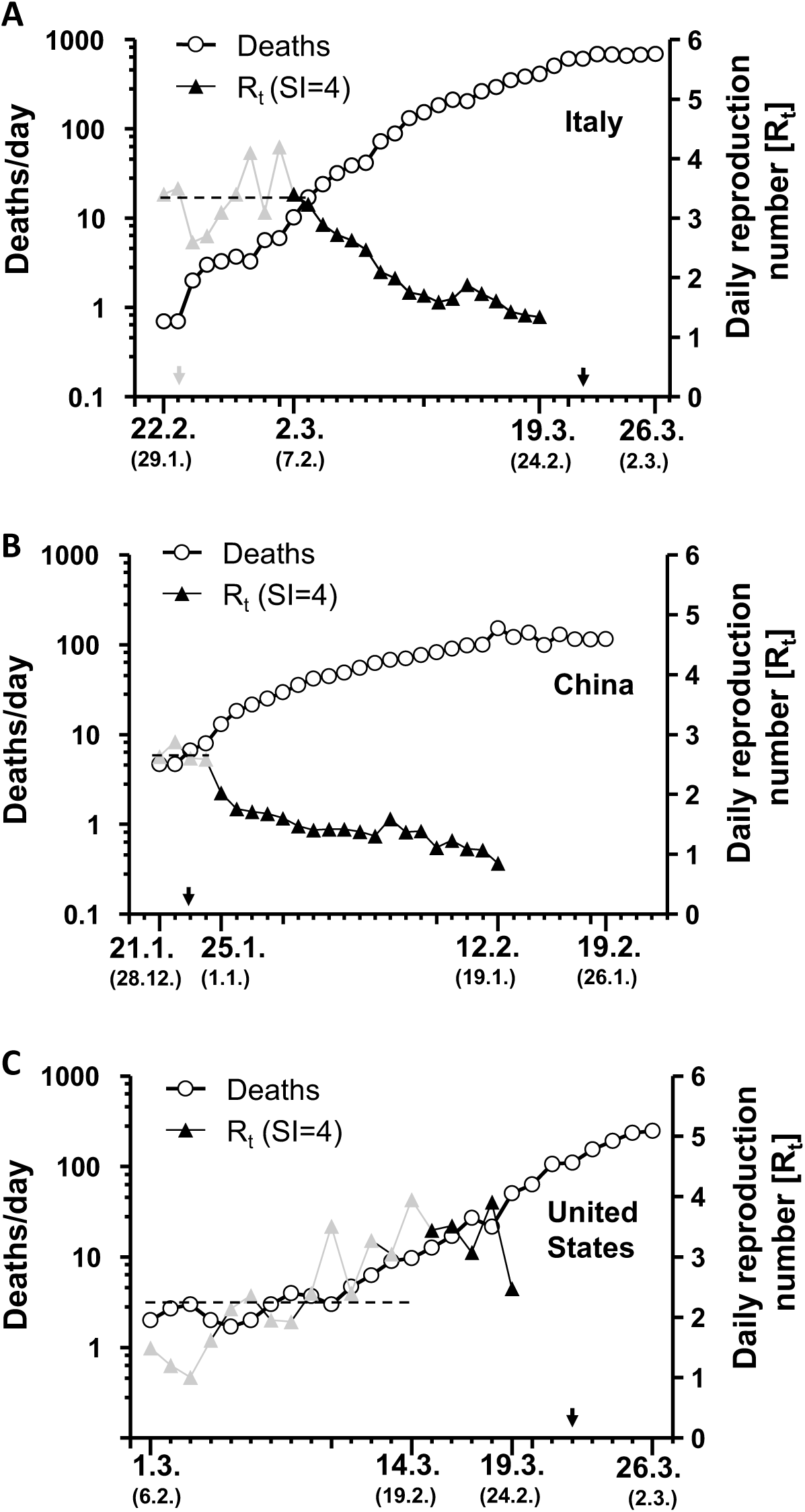
Mean numbers of deaths of the indicated day and the two flanking days in Italy (A), China (B), and the USA (C) are shown for the indicated time periods. The daily reproduction number R_t_ was calculated based on the fold increase during the subsequent 7 days and a serial interval of 4 by taking the nth root of the fold increase with n= fold increase/serial interval. R_t_ values for days with <10 deaths are shown in grey and their mean is indicated by the dashed line. Virus infection is estimated to be acquired on average approximately 24±5 days before the day of death (see text). Thus, the R_t_ values plotted for the day of death reflect the transmission occurring approximately 24 days before (in brackets below the day of death). ↓Start of control measures in Italy. ↓ Shut-down in Italy, Wuhan City, and New York, respectively.

The R_0_ of 3.4 we derive from the rise in early death cases in Italy is higher than the R_0_ of 2.2 reported from the rise in confirmed cases in China ^1^. Our estimate of the R_0_ based on the rise in death cases in China between 21.1.and 24.1.2020 is in the range of 2.7, with high uncertainty due to less than 10 death cases/day (Fig. 1B). Thereafter, the R_t_ declines below 1. For the United States, the R_t_s determined during 1.3. to 14.3.2020 trend to increase to values above 3 (Fig. 1C). The low number of deaths observed during this period may be driven by imported cases rather than autochthonous spread of SARS-CoV2. Thereafter, the mean R_t_ is 3.3 suggesting fast spread of the virus end of February and early March. Since hardly any control measures were implemented in the United States during this time period we consider this an adequate estimate of R_0_.

Thus, our analysis provides evidence that basic epidemiological parameters differ between countries to an extent compromising epidemiological predictions of the pandemic. It also suggests that suppression of spread in Italy and the US may be more difficult to achieve. Although we assume that variations in social behaviour are responsible for the different estimates of R_0_, selection of more rapidly spreading variants of SARS-CoV-2 cannot be excluded. Despite uncertainty in the reliability of the data used and lack of information on possible changes in the effectiveness of registration of COVID-19 deaths during the observation period, our findings should be considered as a working hypothesis demanding further investigations. As the number of deaths rapidly increases worldwide, we encourage more sophisticated modelling of the epidemic based on the dynamics of death cases by experts in the field.

## Data Availability

All data is available and within the manuscript

